# Diastolic Dysfunction in Congenital Heart Disease: A Pressure-Volume Loop Analysis of Borderline Left Heart

**DOI:** 10.64898/2026.01.13.26344073

**Authors:** Nikhil Thatte, Martin Liberman, Peter E. Hammer, Ryan Callahan, Jesse Esch, Michael Farias, Gerald Marx, Rebecca S. Beroukhim, Eric Feins, Sitaram Emani, Sunil J. Ghelani

## Abstract

**Background:** Diastolic dysfunction is a major driver of morbidity in congenital heart disease (CHD), but its mechanisms are poorly understood. It remains unknown whether left ventricular (LV) diastolic dysfunction in borderline left heart patients is caused by a defect in active relaxation or in passive compliance.

**Objectives:** We sought to apply gold-standard pressure-volume (PV) loop analysis to define the primary mechanism of diastolic dysfunction in patients with borderline LV (bLV).

**Methods:** We analyzed invasive PV loop data from pediatric patients with bLV (n =13) and a comparison group of patients with congenitally corrected transposition of the great arteries (ccTGA, presumed no diastolic dysfunction; n=14). PV loops were generated during transient preload reduction and augmentation. Active relaxation was assessed by the isovolumic relaxation time-constant (τ) and maximal rate of pressure decline (dP/dt_min_) indexed to peak-systolic pressure (PSP). Passive compliance was quantified by the chamber stiffness constant (β), derived from the multi-beat end-diastolic PV relationship (EDPVR). To account for size differences, β was (1) calculated using indexed ventricular volumes (iβ), and (2) converted to a dimensionless stiffness constant (β_W_) by multiplying β by ventricular wall volume.

**Results:** Active relaxation measures were similar between groups, indicating normal lusitropy in bLV (τ bLV: 28 [23.5-29.5] vs ccTGA: 24 [21-28] msec, p=0.10; dP/dt_min_/PSP bLV: -10.6 [-11.0, -9.4] vs. ccTGA: -9.9 [-10.5, -8.7] s^-1^, p=0.17). In contrast, patients with bLV had markedly abnormal passive compliance. The EDPVR was shifted sharply upward and to the left, and the stiffness constants were significantly higher in the bLV group (iβ bLV: 0.10 [0.04-0.35] vs. ccTGA: 0.03 [0.02-0.04] m^2^/mL, p=0.003); β_W_ bLV: 4.08 [1.60-7.52] vs. ccTGA: 1.21 [0.95-1.77], p=0.002).

**Conclusions:** The primary mechanism of diastolic dysfunction in bLV is impaired passive ventricular compliance, and not impaired active relaxation. These findings suggest a limited role for lusitropic medications and therapeutic strategies that focus on mitigating ventricular stiffness (such as surgical endocardial fibroelastosis resection) may be more valuable.

**Condensed abstract:** Pressure-volume loop analysis with preload alteration in pediatric congenital heart disease shows that in borderline left heart patients left ventricular diastolic dysfunction is caused by impaired passive compliance, with preserved active relaxation. This mechanistic insight suggests that interventions targeting ventricular stiffness such as surgical endocardial fibroelastosis resection may be more beneficial than the use of lusitropic medications to improve outcomes.

## Introduction

Diastolic dysfunction is a central pathophysiological feature in many forms of congenital heart disease (CHD) and represents a major determinant of clinical outcomes and surgical decision-making.^1–7^ Despite its clinical importance, a precise understanding of the underlying mechanisms in pediatric patients has been limited. Clinical assessment has traditionally relied on non-invasive echocardiographic indices developed for adults with biventricular hearts or on isolated measurements of end-diastolic pressure (EDP). These methods lack the level of detail required to accurately characterize the diastolic properties of the structurally and physiologically unique ventricles found in complex CHD.^2,8^

Pressure-volume (PV) loop analysis is the established gold standard for comprehensively and accurately assessing ventricular diastolic function. By separately studying the active and passive components of diastole, this methodology provides mechanistic insights that are unattainable with other techniques. Active myocardial relaxation (lusitropy), an energy-dependent process, is best quantified by the isovolumic relaxation time constant, τ.^9^ In contrast, passive chamber stiffness, a mechanical property of the ventricle, is defined by the slope of the end-diastolic pressure-volume relationship (EDPVR), characterized by the stiffness constant β.^10^ This methodology has been successfully used in adults to understand and define the syndrome known as heart failure with preserved ejection fraction.^11^ Despite its power, the application of PV loop analysis to systematically study diastolic function in pediatric CHD has been limited, leaving a critical gap in our understanding.

This study sought to address this gap by applying invasive PV loop analysis to a key pediatric population where diastolic function is of particular interest: patients with borderline left hearts being considered for left ventricular (LV) recruitment for biventricular repair vs. Fontan palliation. The presence of endocardial fibroelastosis (EFE), an abnormal thickening of the endocardium, is believed to be a primary contributor to this diastolic impairment.^12,13^ However, it remains unknown whether this dysfunction arises from impaired active relaxation, increased passive stiffness, or both. We hypothesized that in patients with borderline LV (bLV), diastolic dysfunction is predominantly caused by increased passive chamber stiffness rather than by impaired active relaxation. To test this hypothesis, we compared patients with bLV to a comparator cohort of patients with congenitally corrected transposition of the great arteries (ccTGA), a population of similar age and size that routinely undergoes PV loop analysis at our institution for separate clinical indications (analysis of LV preparedness for double switch operation) but typically lacks clinically significant intrinsic diastolic impairment.

## Methods

### Study Design

We performed a retrospective analysis of pediatric patients who underwent cardiac catheterization with PV loop analysis at Boston Children’s Hospital between 2021 and 2025. Patient guardians provided written informed consent for acquisition of PV loop data during the cardiac catheterization. The study protocol was approved by the Institutional Review Board, and the requirement for individual patient consent for this retrospective review was waived.

### Patient Population

Patients were categorized into two groups: (1) bLV group (n=13), and (2) ccTGA comparison group (n=14). The bLV cohort included patients with a mild or moderately hypoplastic LV at various stages of single-ventricle palliation, or post biventricular conversion after initial single ventricle repair. bLV physiology at the time of assessment included stage 1 (n=1), Glenn (n=1), Fontan (n=2), Glenn with an additional source of pulmonary blood flow (n=6), biventricular circulation with LV in systemic position (n=2), LV as the inferior vena cava pump to the lungs (reverse 1.5 ventricle repair; n=1). Nine of 13 (69%) patients had evidence of EFE on cardiac MRI, 3/13 (23%) had evidence of no EFE, and EFE status was unknown in 1/13 (8%). The comparison group consisted of patients with ccTGA and an intact ventricular septum who underwent subpulmonary LV PV loop analysis as part of their clinical evaluation for double switch operation preparedness. These patients had either native LV outflow tract obstruction (n = 3) or had previously undergone main pulmonary artery band placement for LV re-training (n=11). This cohort was selected because of its similarity in age and body size to the bLV group and the typical absence of clinically significant diastolic dysfunction, as the acquisition of normative data from healthy children was not feasible.

### Invasive Hemodynamic Assessment

Under general anesthesia, a micromanometer-tipped conductance catheter (CD Leycom, Netherlands) was placed in the LV. The approach was chosen depending on the underlying anatomy: in ccTGA patients, antegrade via the right internal jugular vein; in bLV patients, retrograde via the femoral artery or antegrade via the femoral vein and across the atrial septum depending on the physiology and at the discretion of the catheterizing physician. Conductance-derived volumes were calibrated with the use of end-systolic and end-diastolic volumes from prior cardiac MRI (n=22) or 3D echocardiography (n=5).^14,15^ To define the EDPVR, a family of PV loops was generated by altering loading conditions through transient inferior vena cava occlusion for preload reduction and a 5 to 10 ml/kg crystalloid bolus or transient atrial septal defect occlusion for preload augmentation. All recordings were performed during breath-holding at end-expiration. Recordings containing ectopic beats or excessive artifact were excluded.

### Pressure-Volume Loop Analysis

PV loop analysis was performed offline using specialized software: ConductNT, CD Leycom, Netherlands.

#### Indices of Active Relaxation

Two metrics were recorded, (1) The time constant of isovolumetric relaxation (τ)^16,17^; higher values indicate worse active relaxation, and (2) the maximum rate of ventricular pressure decline (dP/dt_min_); less negative values indicate worse active relaxation. This metric is load-dependent^18^ so we also report the value of dP/dt_min_ indexed to peak systolic pressure.

#### Indices of Passive Stiffness

The terms compliance and elastance are both used throughout the manuscript – these quantities are simply reciprocals of one another (i.e. elastance = 1/compliance). A stiffer ventricle is one with higher values of diastolic elastance metrics (i.e. ‘less compliant’).

Single-beat end-diastolic chamber elastance (end-diastolic elastance, E_ed_) ^9,19^ was calculated as EDP/end-diastolic volume (EDV); indexed E_ed_ = EDP/EDV indexed to body surface area (BSA).^9,19^ A multi-beat EDPVR was constructed from the family of PV loops obtained from preload alteration by fitting end-diastolic points to the exponential equation P = C * e^βV^, where P is pressure, V is volume, C is a fitting constant, and β is the stiffness constant which represents LV chamber stiffness (Figure 1).^10^ Exponential curve fitting was performed in GraphPad Prism 10 (GraphPad Software, San Diego, CA).

**Figure 1.**
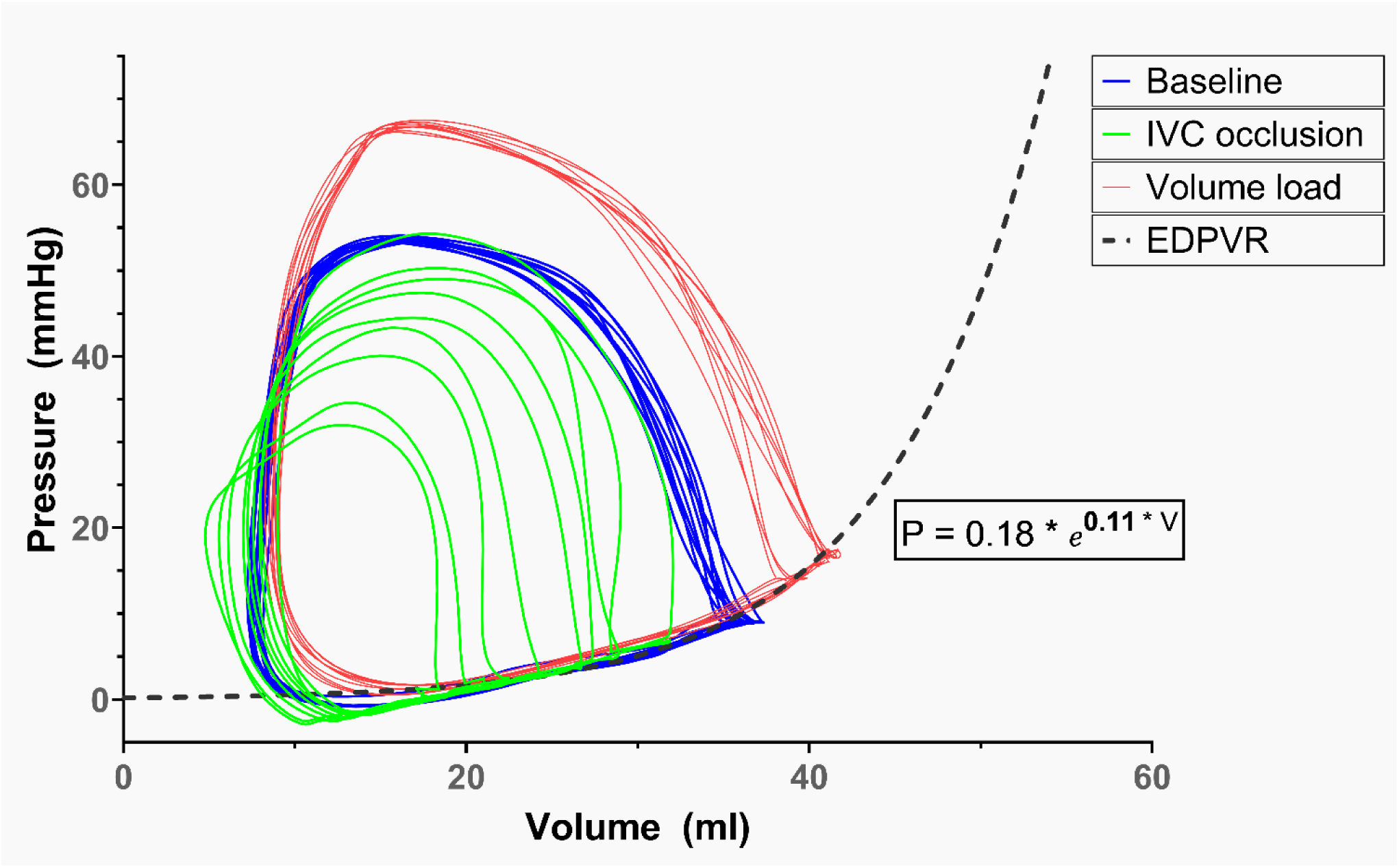
Characterization of the end-diastolic pressure volume relationship (EDPVR). Pressure-volume loops are obtained at baseline (blue loops) and with changes to preload by transient occlusion of the inferior vena cava (green loops) and by administration of a crystalloid volume bolus (red loops). An exponential curve (dashed black line) is plotted to fit the end-diastolic points of the resultant family of loops using the equation 𝑃 = 𝐶 ∗ 𝑒^𝛽𝑉^, where P is pressure, V is volume, C is a fitting constant, and β is the stiffness constant which represents LV chamber stiffness. In this representative patient from the borderline LV group, β = 0.11.

To account for differences in heart and body size, we calculated (1) an indexed stiffness constant, iβ, using ventricular volumes indexed to body surface area (BSA); and (2) a dimensionless chamber stiffness index, βw = β×Vw, where Vw is the ventricular wall volume (see Appendix).^10,20^ Higher values of these stiffness indices represent stiffer ventricles (worse passive compliance). Ventricular capacitance at a pressure of 15 mmHg (V15), another size-independent measure, was also calculated from the EDPVR and indexed to BSA; a lower value indicates a stiffer ventricle (see Appendix).^10^

#### Filling Rates

The peak filling rate (PFR, ml/s) was measured directly from the PV loop data in the baseline condition. It was indexed to the ventricular stroke volume to account for differences in ventricular size and transmitral flow volume.

### Statistical Analysis

Continuous variables were expressed as median and interquartile range. Categorical data were expressed as frequencies and percentages. Fisher’s exact test was used to compare categorical variables, and the Mann-Whitney U test to compare continuous variables. A p-value of <0.05 was considered statistically significant. Analysis was performed using SPSS version 29.0 (IBM Corp, 2022).

## Results

Demographic variables, conventional imaging and catheterization metrics are detailed in Table 1. The groups were similar in age and body size. Indexed LV EDV and LV stroke volume were smaller in the bLV group (indexed LV EDV: bLV 37.8 ml/m^2^ [29.1, 79.7] vs ccTGA 59.6 ml/m^2^ [52.2, 86.1], p=0.029; indexed LV stroke volume: bLV 26.7 ml/m^2^ [14.7, 43.9] vs ccTGA 38.4 ml/m^2^ [33.1, 55.2], p=0.019). LV EF was similar (bLV 62.1% [55.45, 70.3] vs ccTGA 62.15% [55.5, 69.55], p=1). LV end-diastolic pressure was significantly higher in the bLV group at 13 mmHg [8.5, 17.5] vs ccTGA 8 mmHg [6.75, 8.25], p<0.001. Amongst ccTGA patients, median subpulmonary LV pressure was isosystemic (median LV:RV pressure ratio 0.99).

**Table 1.** Patient characteristics.

| Variable | bLV (n=13) | ccTGA (n=14) | p value |
| --- | --- | --- | --- |
| **Demographics** |  |  |  |
| Age at catheterization, years | 4.75 [3.3, 12.88] | 2.9 [1.76, 5.8] | 0.185 |
| Sex, male | 10 (77%) | 11 (79%) | 1 |
| Weight, kg | 17.6 [13.9, 29] | 13.75 [11.15, 18.2] | 0.155 |
| Height, cm | 102 [94, 137.9] | 95.6 [80.1, 114.2] | 0.28 |
| Body surface area, m<sup>2</sup> | 0.71 [0.61, 1.05] | 0.61 [0.51, 0.75] | 0.22 |
| **LV Imaging metrics** |  |  |  |
| Indexed end-diastolic volume, ml/m<sup>2</sup> | 37.8 [29.1, 79.7] | 59.6 [52.2, 86.1] | **0.029** |
| Indexed end-systolic volume, ml/m<sup>2</sup> | 12.04 [8.0, 35.4] | 20.80 [18.7, 36.4] | 0.169 |
| Indexed stroke volume, ml/m<sup>2</sup> | 26.7 [14.7, 43.9] | 38.4 [33.1, 55.2] | **0.019** |
| Ejection fraction, % | 62.1 [55.5, 70.3] | 62.15 [55.5, 69.6] | 1 |
| Indexed LV Mass, g/m<sup>2</sup> | 36.6 [24.3, 48.3] | 43.1 [32.9, 51.5] | 0.28 |
| Mass-to-volume ratio, g/ml | 0.79 [0.6, 0.95] | 0.61 [0.52, 0.77] | 0.054 |
| Ventricular wall volume, ml | 24.99 [16.4, 36.1] | 24.3 [22.0, 29.8] | 0.981 |
| **Catheterization metrics** |  |  |  |
| Baseline heart rate, bpm | 90 [81.5, 107] | 102 [79.25, 109.25] | 0.72 |
| LV peak systolic pressure, mmHg | 101 [90.5, 116] | 80 [61, 106.75] | 0.061 |
| LV end-systolic pressure, mmHg | 89 [85.5, 104.5] | 59 [49, 89.25] | **0.012** |
| LV end-diastolic pressure, mmHg | 13 [8.5, 17.5] | 8 [6.75, 8.25] | **<.001** |
| LV:RV pressure ratio | n/a | 0.99 [0.72, 1.24] | n/a |

Values expressed as n (%) or Median [IQR].
Abbreviations: bLV, borderline left ventricle; ccTGA, congenitally corrected transposition of the great arteries; RV, right ventricle

### Active Relaxation

The metrics of active relaxation were similar between the two groups (Table 2). The time constant of isovolumetric relaxation τ was not significantly different between the bLV group and the ccTGA group (τ bLV 28 msec [23.5, 29.5] vs ccTGA 24 msec [21, 28], p=0.105). The uncorrected dP/dt_min_ was more negative in the bLV group (indicating better active relaxation), but this difference did not persist after indexing to peak systolic pressure - uncorrected dP/dt_min_: bLV -1040 mmHg/s [-1239, -897] vs ccTGA -781 mmHg/s [-913.75, -572.75], p = 0.008; dP/dt_min_/peak systolic pressure: bLV -10.55 s^-1^ [-11.03, -9.42] vs ccTGA -9.87 s^-1^ [-10.45, -8.67], p = 0.169.

**Table 2.** Metrics of left ventricular active relaxation.

| Variable | bLV (n=13) | ccTGA (n=14) | p value |
| --- | --- | --- | --- |
| Tau ($\tau$), msec | 28 [23.5, 29.5] | 24 [21, 28] | 0.105 |
| dP/dt<sub>min</sub>, mmHg/s | -1040 [-1239, -897] | -781 [-913.75, -572.75] | **0.008** |
| dP/dt<sub>min</sub>/PSP, s<sup>-1</sup> | -10.55 [-11.03, -9.42] | -9.87 [-10.45, -8.67] | 0.169 |

Values expressed as Median [IQR]. Abbreviations: bLV, borderline left ventricle; ccTGA, congenitally corrected transposition of the great arteries; PSP, peak systolic pressure

### Passive Compliance

In contrast to active relaxation, passive compliance was markedly different in patients with bLV compared to those with ccTGA (Table 3). Single-beat end-diastolic elastance E_ed_ as well as indexed E_ed_ were significantly higher in the bLV group (indexed E_ed_ bLV 0.32 mmHg/ml/m^2^ [0.20, 0.52] vs ccTGA 0.11 mmHg/ml/m^2^ [0.08, 0.15], p<0.001). The chamber stiffness constant β, the indexed constant iβ, and the dimensionless index βw were all approximately threefold higher in the bLV group, indicating significantly worse passive compliance (chamber stiffness index, βw (dimensionless) bLV 4.08 [1.6, 7.52] vs ccTGA 1.21 [0.95, 1.77], p = 0.002; indexed stiffness constant iβ, bLV 0.10 m^2^/mL [0.04, 0.35] vs ccTGA 0.03 m^2^/mL [0.02, 0.04], p = 0.003). Figure 2 displays the EDPVR of each individual patient, with a median curve for each group shown in bold. Compared to the ccTGA patients (green lines), the EDPVRs for the bLV patients (red lines) are generally shifted more upwards and to the left indicating stiffer ventricles in the bLV group. The median ventricular capacitance at 15 mmHg (V15) was significantly lower in the bLV group compared to the ccTGA group, again indicating a stiff, noncompliant chamber (bLV 39.15 ml/m^2^ [28.55, 74.54] vs ccTGA 95.85 ml/m^2^ [65.43, 127.57], p=0.001).

**Figure 2.**
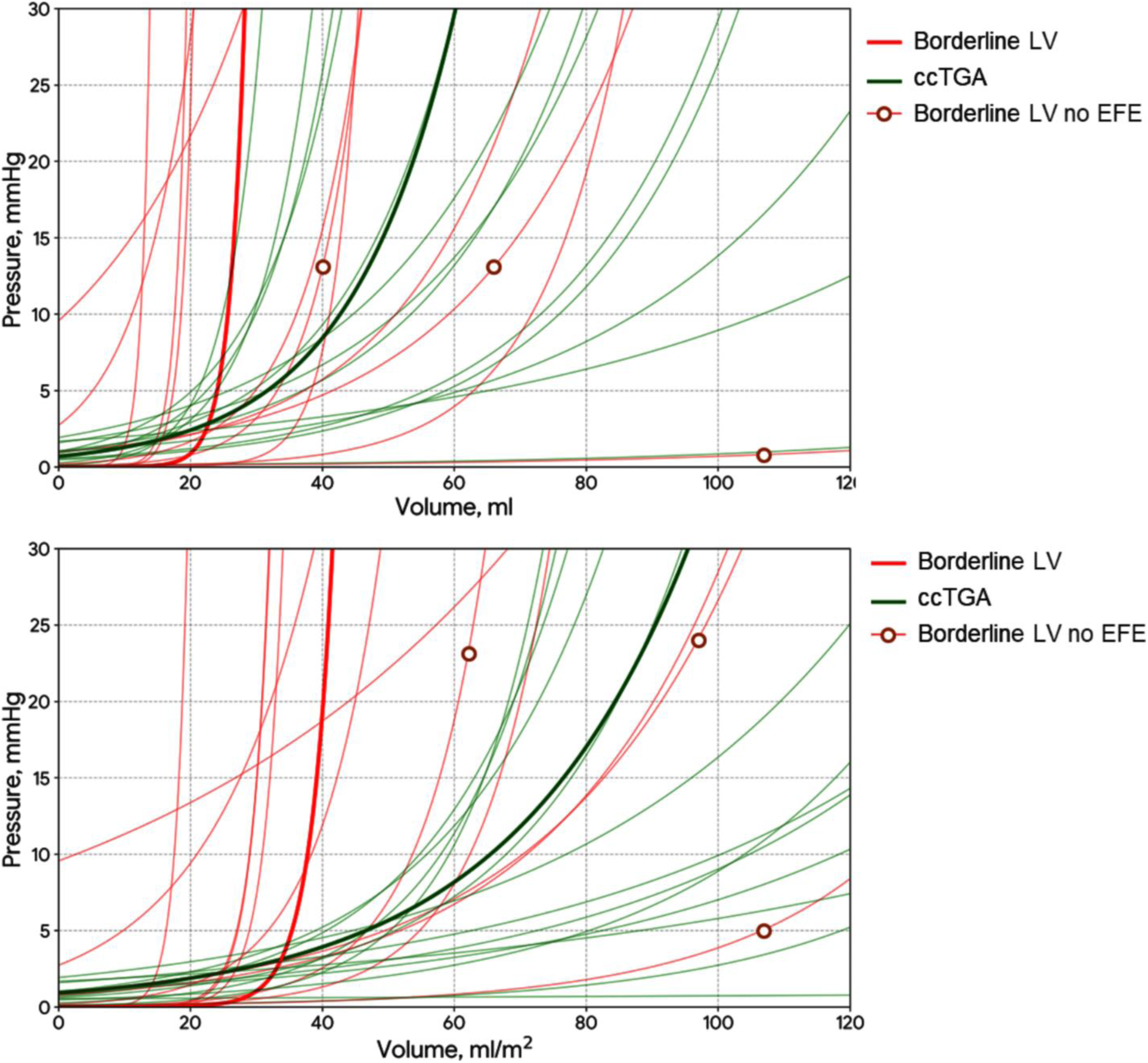
The end-diastolic pressure-volume relationships (EDPVR) of all patients is displayed. Each individual patient is displayed as a single line (red – borderline left ventricle (bLV), green – congenitally corrected transposition of the great arteries (ccTGA)). A representative median curve is shown in bold for each group. The EDPVRs for patients with bLV are steeper, shifted up and to the left, compared to ccTGA patients. The three red lines marked with circles indicate bLV patients with no endocardial fibroelastosis (EFE); these patients had relatively flatter EDPVRs than the remaining bLV patients. Top panel: EDPVRs plotted using unindexed ventricular volumes. Bottom panel: EDPVRs plotted using ventricular volumes indexed to body surface area.

**Table 3.** Metrics of left ventricular passive compliance and filling rates.

| Variable | bLV (n=13) | ccTGA (n=14) | p value |
| --- | --- | --- | --- |
| Curve fitting constant, C | 0.05 [0, 0.87] | 0.79 [0.42, 1.19] | **0.025** |
| Single beat end-diastolic elastance (E<sub>ed</sub>), mmHg/ml | 0.31 [0.21, 0.87] | 0.21 [0.11, 0.29] | **0.012** |
| Indexed single beat end-diastolic elastance (indexed E<sub>ed</sub>), mmHg/ml/m<sup>2</sup> | 0.32 [0.20, 0.52] | 0.11 [0.08, 0.15] | **<0.001** |
| Stiffness constant $\beta$, ml<sup>-1</sup> | 0.12 [0.05, 0.51] | 0.04 [0.03, 0.08] | **0.017** |
| Chamber stiffness index, $\beta_w$ | 4.08 [1.6, 7.52] | 1.21 [0.95, 1.77] | **0.002** |
| Indexed Stiffness Constant iβ, m<sup>2</sup>/mL | 0.10 [0.04, 0.35] | 0.03 [0.02, 0.04] | **0.003** |
| Indexed V15, ml/m<sup>2</sup> | 39.15 [28.55, 74.54] | 95.85 [65.43, 127.57] | **0.001** |
| PFR, ml/s | 176 [88, 316.5] | 229.5 [168.75, 352.75] | 0.259 |
| PFR/stroke volume, s<sup>-1</sup> | 8 [7.37, 9.41] | 9.02 [7.52, 10.5] | 0.402 |

Values expressed as Median [IQR]. Abbreviations: bLV, borderline left ventricle; ccTGA, congenitally corrected transposition of the great arteries; PFR, peak filling rate.

### Role of EFE

Within the bLV group, three patients had no EFE on cardiac MRI. These three patients had more rightward-downward shifted (flatter) EDPVRs indicating better compliance (Figure 2, three lines marked with circles) compared to the bLV patients who had evidence of EFE. The sample size was inadequate to perform statistical comparisons between these bLV subgroups.

### Filling Rates

There were no differences between the groups in peak filling rate, both when compared for absolute volumes and when indexed to stroke volume (Table 3).

## Discussion

This study, the first to our knowledge to systematically apply invasive pressure-volume loop analysis to characterize diastolic function in a pediatric CHD cohort, demonstrates that this methodology is feasible and can yield mechanistic data in children with complex cardiac anatomy and physiology. The study shows that LV diastolic function in patients with bLV is characterized by a profound impairment in passive ventricular compliance, while active relaxation is preserved. These results support the hypothesis that the pathophysiology of diastolic dysfunction in bLV is primarily a problem of increased chamber stiffness.

The distinction between impaired relaxation and increased stiffness has important clinical implications. Active relaxation is an ATP-dependent process related to calcium handling, which can be a target for lusitropic medical therapies such as milrinone. Our finding that τ and dP/dt_min_ are preserved in bLV suggests that such therapies may offer limited benefit. Instead, the markedly increased passive stiffness points to a structural cause, such as the myocardial hypertrophy and extensive EFE commonly observed in this condition. This finding provides a mechanistic rationale for therapeutic strategies that directly target ventricular stiffness. The surgical approaches adopted toward this goal in borderline left heart patients include resection of EFE and relief of any residual LV outflow tract obstruction (which leads to ongoing hypertrophy and increases the risk of recurrence of EFE).^21–25^ Medical therapies for LV diastolic dysfunction related to increased ventricular stiffness have historically been limited, but recent studies suggest a potential role for drugs such as sodium-glucose cotransporter-2 (SGLT2) inhibitors^25–28^ and statins^29^ though these studies were not performed in CHD patients.

PV loop analysis overcomes some of the limitations of conventional methods for assessing diastolic function in CHD. Echocardiographic indices are often confounded by loading conditions, heart rate, and atypical chamber geometry. Similarly, a single elevated measurement of EDP, while informative, cannot distinguish between a stiff ventricle and a compliant ventricle that is volume or pressure overloaded. Conversely, a reassuringly normal EDP may be due to an underloaded ventricle which could be unmasked as having severe diastolic dysfunction after a biventricular repair. By generating a family of loops under varying loading conditions, we defined the intrinsic end-diastolic pressure-volume relationship, allowing for an unambiguous quantification of LV diastolic function. The successful application of this technique in two distinct CHD populations demonstrates its potential to advance our understanding and guide management across a wide spectrum of congenital heart defects.

Our study also shows that not all patients with bLV have equally stiff ventricles. Some bLV patients, particularly those without EFE (Figure 2, lines marked with circles), had EDPVRs that were significantly flatter (rightward and downward) than others. Therefore, it is important to study each patient individually to determine their ventricular stiffness, response to volume loading, and suitability for using the ventricle in any proposed future circulation.

## Limitations

Several limitations of this study warrant consideration. First, the sample size is modest. However, it represents the largest dataset of invasively acquired PV loop data with preload alteration in a pediatric congenital heart disease cohort to date.

Second, our comparison group consisted of patients with ccTGA with subpulmonary LV who are not truly normal and may have subtle diastolic abnormalities of their own due to the presence of a main pulmonary artery band. However, this population represents a practical and ethically appropriate control group, as they have complex anatomy and physiology requiring invasive pre-operative hemodynamic assessment but do not typically suffer from the intrinsic myocardial and endocardial stiffness abnormalities that define bLV. The striking differences observed between the groups, despite any subtle abnormalities in the ccTGA cohort, only strengthen our conclusions regarding the severity of impaired compliance in bLV.

Ventricular volumetry by 3D echocardiography has been shown to correlate well with volumes derived from cardiac MRI when performed at an experienced echocardiography laboratory.^14,15^ However, the use of 3D echocardiography-derived LV volumes for calibration of the conductance catheter may introduce a source of error when compared to cardiac MRI-derived volumes which represent the bulk of the patients in our cohort.

Conductance-catheter based recording of PV loops can be technically challenging. Some of these challenges are exacerbated in pediatric patients with congenital heart disease. To acquire analyzable data, the ventricle must have a minimal (end-systolic) long-axis length of at least 33 mm which excludes patients with the most hypoplastic LVs. In patients in whom there is LV hypertrophy with free wall-to-septum contact in systole, the PV loop tracings can contain significant artifact precluding their use for analysis. There is an institutional learning curve to be able to efficiently and accurately collect and analyze PV loop data. Finally, even with an ideal data set, the analysis of the data involves both mathematical and physiological assumptions.

Careful understanding of these both during data acquisition as well as analysis is necessary to minimize errors in interpretation. Despite these challenges, PV loop-based study of diastolic function remains our best method for studying this complex physiologic process.

## Future Directions

Although our data suggest that bLV patients with EFE have worse LV compliance than those without EFE, the sample size was not adequate to definitively demonstrate this. Additionally, the study does not evaluate the impact of surgical EFE resection on ventricular stiffness. Future studies can address this question by evaluating LV diastolic function in a cohort of borderline left heart patients before and after EFE resection. We continue to seek more accurate non-invasive techniques to estimate the invasively obtained PV-loop derived metrics of diastolic function in these patients, and hope to do so in the future using the invasively acquired data in the present study as gold-standard.

## Conclusions

Systematic invasive PV loop analysis is a feasible tool for providing mechanistic insights into diastolic function in complex pediatric CHD. The primary mechanism of DD in bLV is impaired passive ventricular compliance, and not impaired active relaxation. These findings suggest that therapeutic strategies that focus on mitigating ventricular stiffness (such as surgical EFE resection and relief of LV outflow tract obstruction, and medications such as SGLT2 inhibitors) may be more valuable to improve outcomes, while there may be a limited role for lusitropic medications.

## Appendix

1. Ventricular wall volume (Vw) calculation: Vw is the volume of the myocardial shell of the LV. It represents the difference between epicardial volume and endocardial volume, which are both routinely measured by cardiac MRI or 3D echocardiography to calculate LV mass. LV mass is calculated from the LV wall volume using an assumed myocardial density of 1.055 g/ml, using the standard relationship Myocardial Density = LV Wall Mass/LV Wall Volume (Vw). Therefore, Vw can be easily calculated from LV mass as follows: Vw (in ml) = LV Mass (in grams)/1.055.
2. Ventricular capacitance at 15 mmHg (V15): Ventricular capacitance is defined as the ventricular volume at a specified pressure. ^10^ The value is obtained by first estimating the patient’s exponential curve fit equation P = C * e^βV^ and then solving for P = 15. The resultant value of V is the V15. A smaller value of V15 indicates a stiffer ventricle which can accommodate less volume to reach this specified end-diastolic pressure.

## Data Availability

Available upon reasonable request

## Notes

### Competing Interest Statement

The authors have declared no competing interest.

### Funding Statement

No funding disclosures

### Author Declarations

Boston Childrens Hospital Institutional Review Board

